# KNOWLEDGE AND UPTAKE OF TETANUS TOXOID VACCINE AND ASSOCIATED FACTORS AMONG REPRODUCTIVE AGE GROUP WOMEN IN HAYK TOWN SOUTH WOLLO, ETHIOPIA, CROSS-SECTIONAL STUDY

**DOI:** 10.1101/2022.12.20.22283731

**Authors:** Tiruset Gelaw, Sindu Ayalew, Kassaw eyene

**Affiliations:** Department of Midwifery Collage of Medicine and Health Sciences, Wollo University, Dessie, Ethiopia; Department of Midwifery Collage of Medicine and Health Sciences, Arba Minch University, Arba Minch, Ethiopia

**Keywords:** Tetanus toxoid, reproductive age group, complete dose, Ethiopia

## Abstract

**Background:** Tetanus is an acute, often fatal, disease caused by an exotoxin and highly potent neurotoxin, tetanospasm, Tetanus is a preventable disease by tetanus toxoid immunization, which is usually given to the reproductive women at the age between (15-44) years in order to protect both mother and newborn in order to prevent maternal and neonatal tetanus For lifelong protection from tetanus a woman needs a total of five tetanus toxoid doses

**Objective:** To assess the level of knowledge uptake and associated factors of completing tetanus toxoid vaccine among reproductive age women in Hayk town, SouthWollo, Amhara, Ethiopia in 2020.

**Methods and materials:** A community based cross-sectional study conducted in Hayk town, South Wollo Amhara, Ethiopia, from November to December on 2020. Data collected using structured and pretested questionnaire, entered in to Epi Data version 3.1 and exported to statistical package for social science version 25.0 for analysis.

**Result:** Magnitude of uptake of TT2 immunization at Hayk town was 71.2%, have taken at least two doses of TT. But only 35(8.5%) had completed 5 doses of TT according to EPI schedule. Nearly half of 182(44%) of women in the reproductive age group had knowledge on complete TT5 immunization. Educational status, having sort of info on TT5 and knowledge about TT are significantly associated factors for full dose of TT according to EPI schedule.

**Conclusion:** Fair proportions of women had uptake of TT2, but very low proportion for complete dose of TT5 according to expanded program on Immunization. Most of study participants have inadequate knowledge on TT vaccine. Educational status, having sort of info on TT5 and knowledge about TT are significantly associated factors for completing full dose of TT according to EPI schedule.

## Background

Tetanus is caused by toxigenic Tetanus is a very highly dangerous fatal disease with a mortality rate about 35%, and a proximal (309000) deaths occurs due to maternal or neonatal tetanus which reflect a triple failure of public health because of defect in routine immunization program during pregnancy period, poor ante-natal care health services, increase home delivery, and unhygienic delivery(1).

The World Health Organization estimated that there were 34 000 neonatal tetanus deaths worldwide in 2015(2). Women die every year due to maternal tetanus that is responsible for more than 5% of maternal deaths and 30, 000 women affected by the tetanus disease(3). The African Region has the highest burden of tetanus with 44 % of the global disability adjusted life years(4)

Tetanus is a preventable disease by tetanus toxoid immunization. Tetanus Toxoid vaccine is usually given to the reproductive women at the age between (15-44) years in order to protect both mother and newborn in order to prevent maternal and neonatal tetanus (5). The tetanus toxoid is a vaccine used in the management and treatment of tetanus. It is highly effective when used prophylactically in cases when dealing with metal injury wound care and management(6).

A completed primary course of TT immunization induces protective immunity that persists for at least 10 to 30 years; persons who have not completed a primary series may require TT(7) since the vaccine economic burden is high,donor support continues to enable the country’s effort to fulfill immunization coverage(8).

In view of maternal and neonatal tetanus (8), TTI must be fully-expanded in women reproductive age-group. TTI also needs to be promoted at injuries during sports or other activities like military training camps. Tetanus-prone wounds not only should receive passive immunization, but emphasis should also be given to completing TTI schedules.

Even though there is a good progress towards maternal and neonatal tetanus elimination the disease remains an important global public health problem, particularly in settings with high neonatal mortality and among some of the poorest countries. Ethiopian government focuses on elimination of MNT and improving maternal health. Ethiopia is one of the countries which have not fully achieved the complete elimination of MNT due to the low TT coverage achieved(8).

Most pregnant women visit ANC at a late time-point of their pregnancy when TTI cannot provide protection(9). Besides, most default from receiving a postpartum dose which would be protective for further pregnancies. Thus this study assesses the knowledge and uptake of complete TT vaccine and associated factors among childbearing age women in Hayk town, South Wollo, Amhara, Ethiopia.

## Materials and Methods

### Study settings, design and population

Community based cross-sectional study was employed in Hayk town, south Wollo Zone, from November to December on 2020. Hayk town, which is located in South Wollo zone, Amhara region, 430 Km away from, Addis Ababa the capital city of Ethiopia. The town has an estimated total population of 14,319 of whom 7, 093 are women, of them 3200 are in age group, (25-45). Most of residence uses agriculture as means of income. The town is known as tourism site with two lakes (Logo haik and Ardebo) and Haik Stefano’s Monastery. The town has one governmental hospital, 1 health center, 2 private pharmacies (10)

### Population

#### Source population

The source population was all child bearing age women who are living in Hayk town

#### Study population

All women in the reproductive age group who were avail in the study period.

### Eligibility criteria

#### Inclusion criteria

All women whose age greater than or equal to18 years old and women who gave birth at least once before two years was included.

#### Exclusion criteria

All reproductive age women who are critically ill were excluded.

### Sample size determination and sampling technique

#### Sample size determination

Different sample sizes are calculated based on the objectives and then by comparing those calculated sample sizes, the largest sample size will be used for this study. The sample size for the first specific objective is determined using single proportion formula by assuming that the prevalence of TT2+ is 51.8 % result of Errer Eastern Ethiopia with 0.05 marginal errors and 95% confidence level of certainty

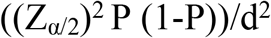

Where n = sample size

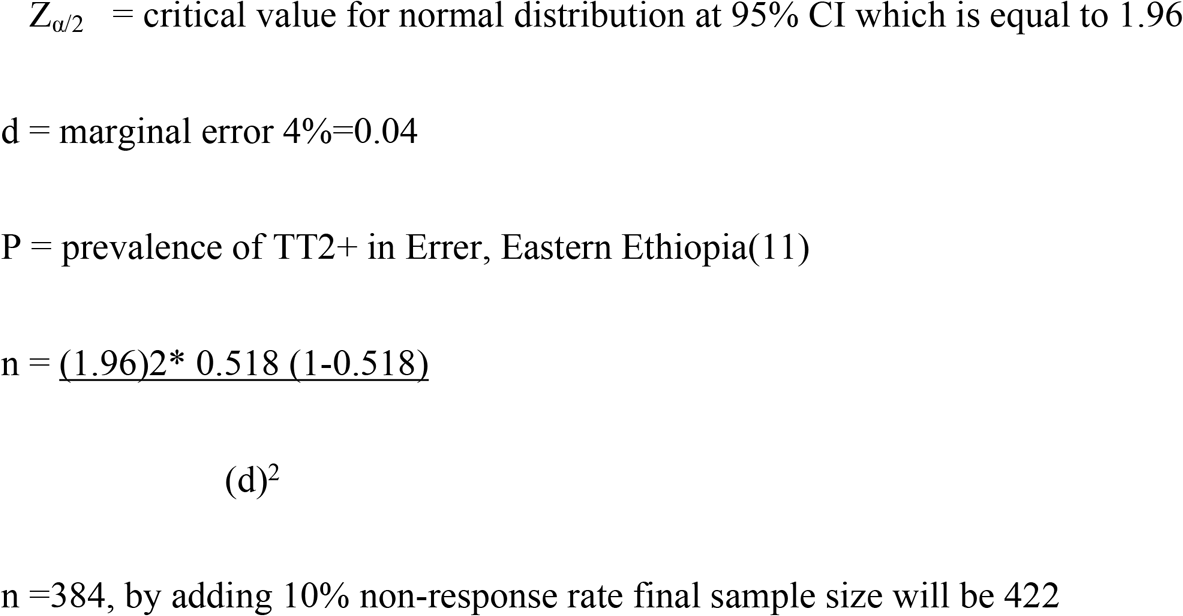

#### Sampling technique

The study participants were selected by systematic random sampling technique one in every 8 women. The first participant was selected by lottery method. The k interval was 3200/422=7.6=8, every k (8 intervals) woman was interviewed.

### Study variables

#### Dependent variable

- Uptake of Tetanus Toxoid vaccine

#### Independent variables

##### Socio demographic characteristics

- Age
- Marital status
- Income/ employment
- Occupation
- Educational status

**Obstetric factors** - Gravida/ Para

Knowledge of Tetanus Tetanus vaccine completion

Uptake of TT vaccine

### Operational Definitions

Uptake: was measured if the mother received≥ TT2 doses as the first dose is only used for the sake of boosting immunity.

Complete TT-taking full dose (6 dose in primary series of TTCV or 5 dose in reproductive age of TT in appropriate schedule

Vaccinated: by card only is when the mother had doses documented on the immunization card (12)

Vaccinated: by card plus history is when both documentation on the immunization card of the mother and doses reported by the mother were considered (12).

Knowledge ability: is measured the respondents correctly answer>3 out of 5 knowledge questions about tetanus toxoid vaccination (11).

### Data collection method and instrument

Data was collected using semi structured and pretested questionnaire by face to face interview technique. Six midwives will collect the data after taking training. The questionnaire includes socio demographic data, obstetric history, knowledge and uptake of TT vaccine.

### Data quality assurance and analysis

#### Data quality assurance

Data quality was assured before, during and after data collection. Before data collection, the questionnaire was first prepared in English language and translated into Amharic and then translated back to English in order to ensure its consistency. Prior to actual data collection pretest was done on 5% of the sample size in 21 to determine its clarity, adequacy, and efficiency of the questionnaire. One day training was given for data collectors and supervisors. Corrections and modifications may be taken according to the result. Data was collected by six trained midwives, in each health facility and each filled questionnaire was checked daily for completeness and accuracy.

During data collection period, study participants were informed about the purpose of data collection and importance of the study to generate quality data. The collected data was checked for completeness and consistencies by trained supervisors and investigator through close follow up and immediate action was taken accordingly. After data collection, the collected data was rechecked for completeness and consistencies by investigator.

#### Data processing and analysis

Data was entered in to Epi data version 3.1 after checking their completeness and exported to statistical package for social science SPSS version 25.0 for analysis. Descriptive statistical data analyzed and results presented by table, graphs and pie charts. Binary Logistic regression used to find out association between explanatory and response variables. Multiple Logistic regressions were used to find out association between dependent and independent variables. P-value less than 0.25 from binary logistic regression were considered for the multivariable logistic regression model and the model goodness of fit was tested using Hosmer and Lemeshow test and strength of association was evaluated using odds ratio at 95% confidence interval and P-values than 0.05 was considered to declare significant associations.

##### Ethical clearance

Ethical clearance and permission was obtained from the Ethical review committee of Wollo University, college of medicine and health Science and the respective health institutions before the data collection process. All the study participants were informed about the objectives and importance of the study. Also, the study subjects were informed that information is coded and used only for research purpose and then verbal consent was taken. The respondents had full right to refuse participation or terminate their involvement at any point during the interview. The confidenciality of study participant information was ensured.

## Results

### Socio demographic status of the respondents

Table 1 A total of 414 participated in the study making a response rate of 98.1%. Almost all of participants are married, and Amhara in Ethnicity (400(96.6%) (Table 1)

**Table 1.** sociodemographic status of study participants, Haik,Wollo, Ethiopia, 2020.

| Variables |  | Frequency | percent |
| --- | --- | --- | --- |
| Marital status | Married | 390 | 94.2 |
|  | Single | 5 | 1.2 |
|  | Divorced | 11 | 2.7 |
|  | Widowed | 8 | 1.9 |
|  | Total | 414 | 100 |
| Religion | Orthodox<br>Christian | 148 | 35.7 |
|  | Muslim | 259 | 59.4 |
|  | Protestant | 7 | 1.7 |
|  | Total | 414 | 100 |
| Age | <20 | 3 | .7 |
|  | 20-24 | 30 | 7.2 |
|  | 25-29 | 53 | 12.8 |
|  | 30-34 | 147 | 35.5 |
|  | 35-39 | 114 | 27.5 |
|  | >-40 | 67 | 16.2 |
|  | Total | 414 | 100.0 |
| Ethnicity | Amhara | 400 | 96.6 |
|  | Tigre | 13 | 3.1 |
|  | Oromo | 1 | 0.2 |
|  | Total | 414 | 100.0 |
| Educational status of women | No formal education | 62 | 15 |
|  | Primary education | 122 | 29.5 |
|  | Secondary education | 145 | 35.5 |
|  | Collage and above | 83 | 20 |
|  | Total | 414 | 100 |
| Occupation of women | Employee | 72 | 17.4 |
|  | Housewife | 246 | 59.4 |
|  | Merchant | 90 | 21.7 |
|  | Other | 6 | 1.4 |
|  | Total | 414 | 100 |
| Educational status of husband | No formal education | 23 | 5.9 |
|  | Primary education | 100 | 25.6 |
|  | Secondary education | 122 | 31.3 |
|  | Collage and above | 145 | 37.2 |
|  | Total | 390 | 100.0 |
| Occupation of husband | Employee | 123 | 31.5 |
|  | Merchant | 239 | 61.3 |
|  | Farmer | 18 | 4.6 |
|  | Other | 10 | 2.6 |
|  | Total | 390 | 100 |

### Obstetrics and other health related issues

Most of 384(84.1%) the participants had antenatal care visit in their past pregnancy and most of them gave birth at health institute. Nearly half of the participants had nearby health facility which is average walking time of 15-30 minutes from their home.(Table 2)

**Table 2.**
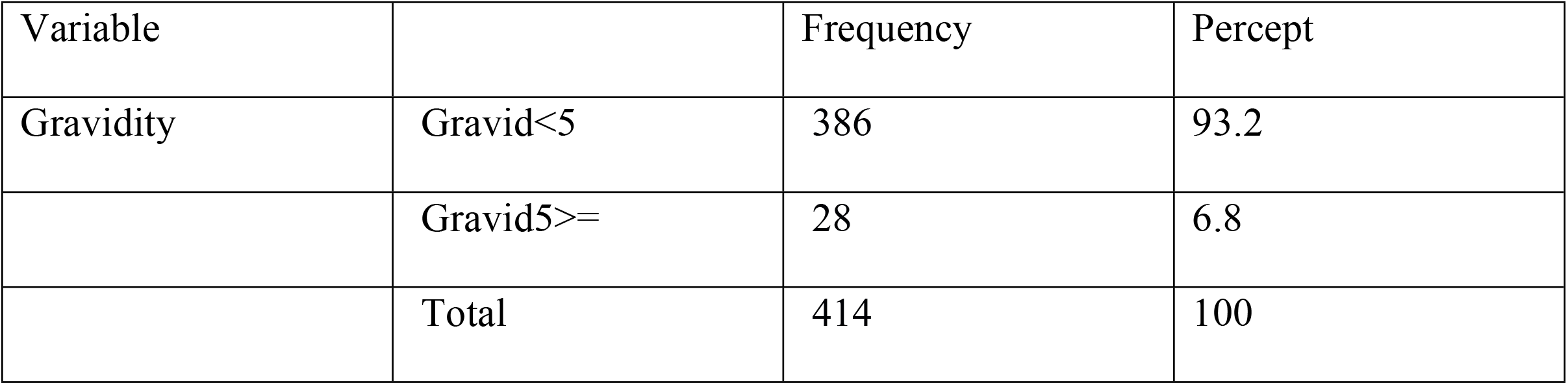
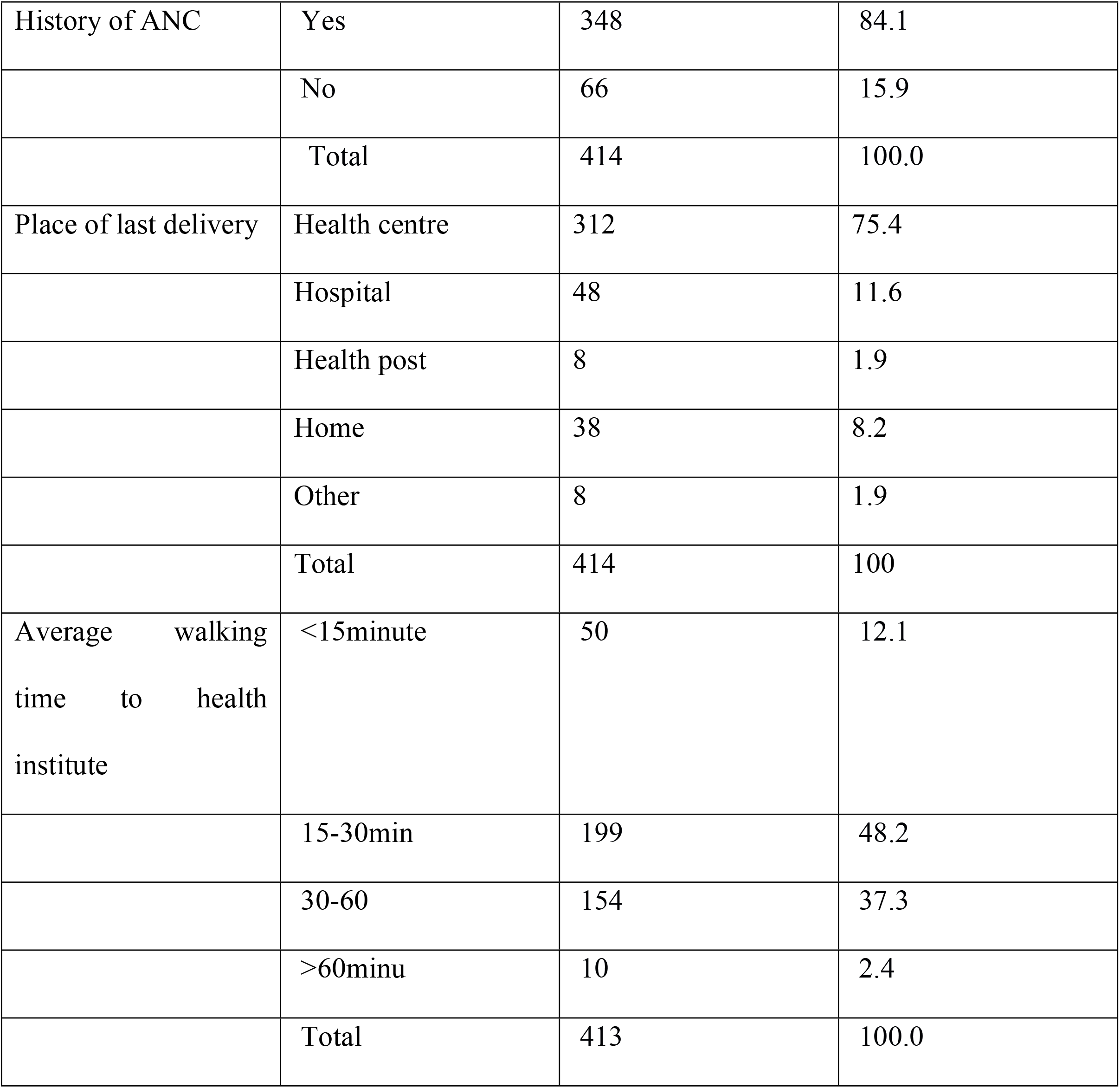
Obstetric history and other health related responses of women tudy participants,Haik,Wollo, Ethiopia, 2020

| Variable |  | Frequency | Percept |
| --- | --- | --- | --- |
| Gravidity | Gravid<5 | 386 | 93.2 |
|  | Gravid5>= | 28 | 6.8 |
|  | Total | 414 | 100 |
| History of ANC | Yes | 348 | 84.1 |
|  | No | 66 | 15.9 |
|  | Total | 414 | 100.0 |
| Place of last delivery | Health centre | 312 | 75.4 |
|  | Hospital | 48 | 11.6 |
|  | Health post | 8 | 1.9 |
|  | Home | 38 | 8.2 |
|  | Other | 8 | 1.9 |
|  | Total | 414 | 100 |
| Average walking time to health institute | <15minute | 50 | 12.1 |
|  | 15-30min | 199 | 48.2 |
|  | 30-60 | 154 | 37.3 |
|  | >60minu | 10 | 2.4 |
|  | Total | 413 | 100.0 |

Other-on the transport, private clinic

### Knowledge and Uptake of TT vaccine

Table 3 showed that two hundred ninety eight (71.2%) of the study participants have vaccinated at least TT2 vaccine and of them only 11.5%(valid percent) of women took TT complete dose according to EPI schedule correctly. One hundred eight two (44%) of study participant had good knowledge on TTT vaccine(Table 3).

**Table 3.**
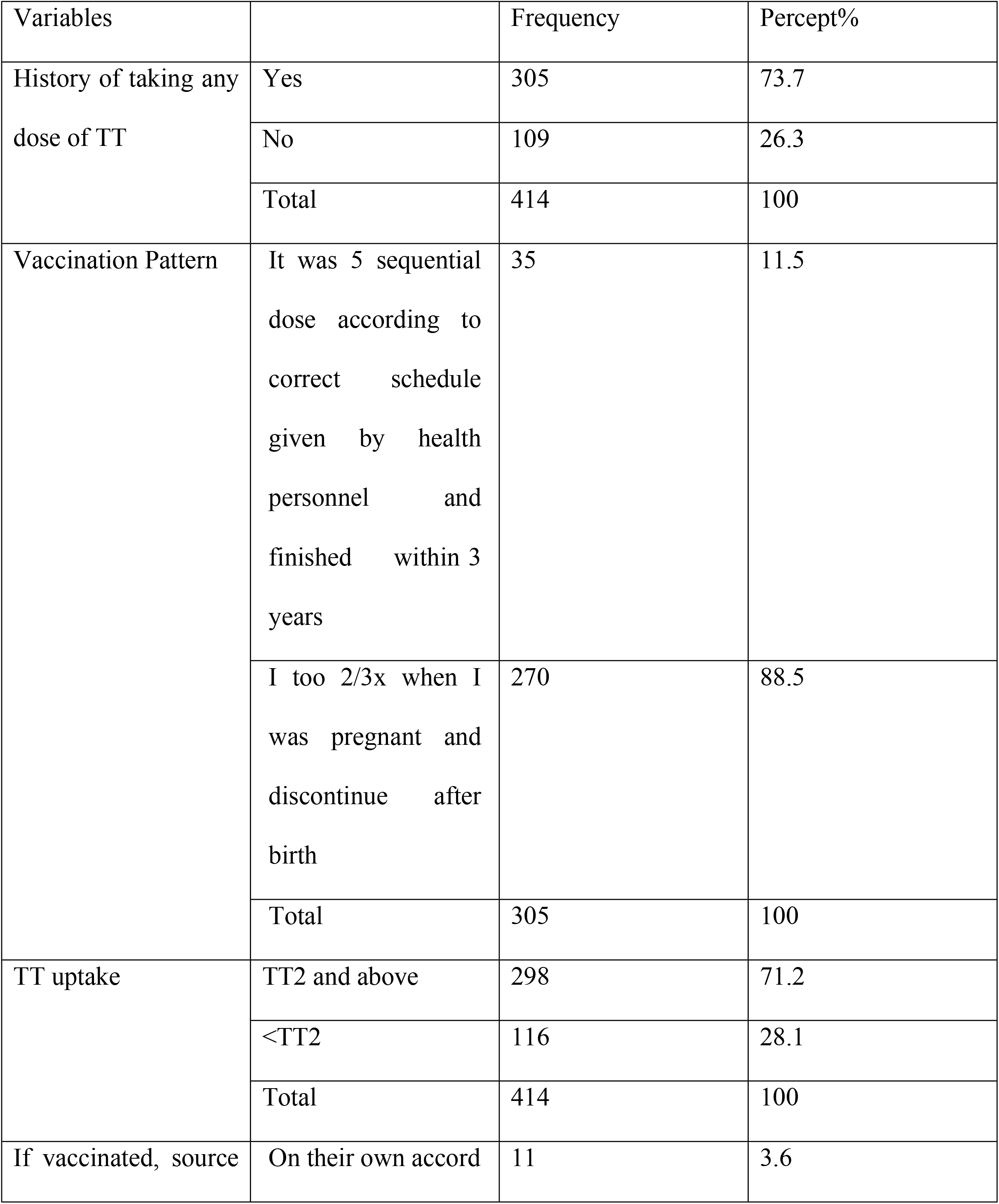
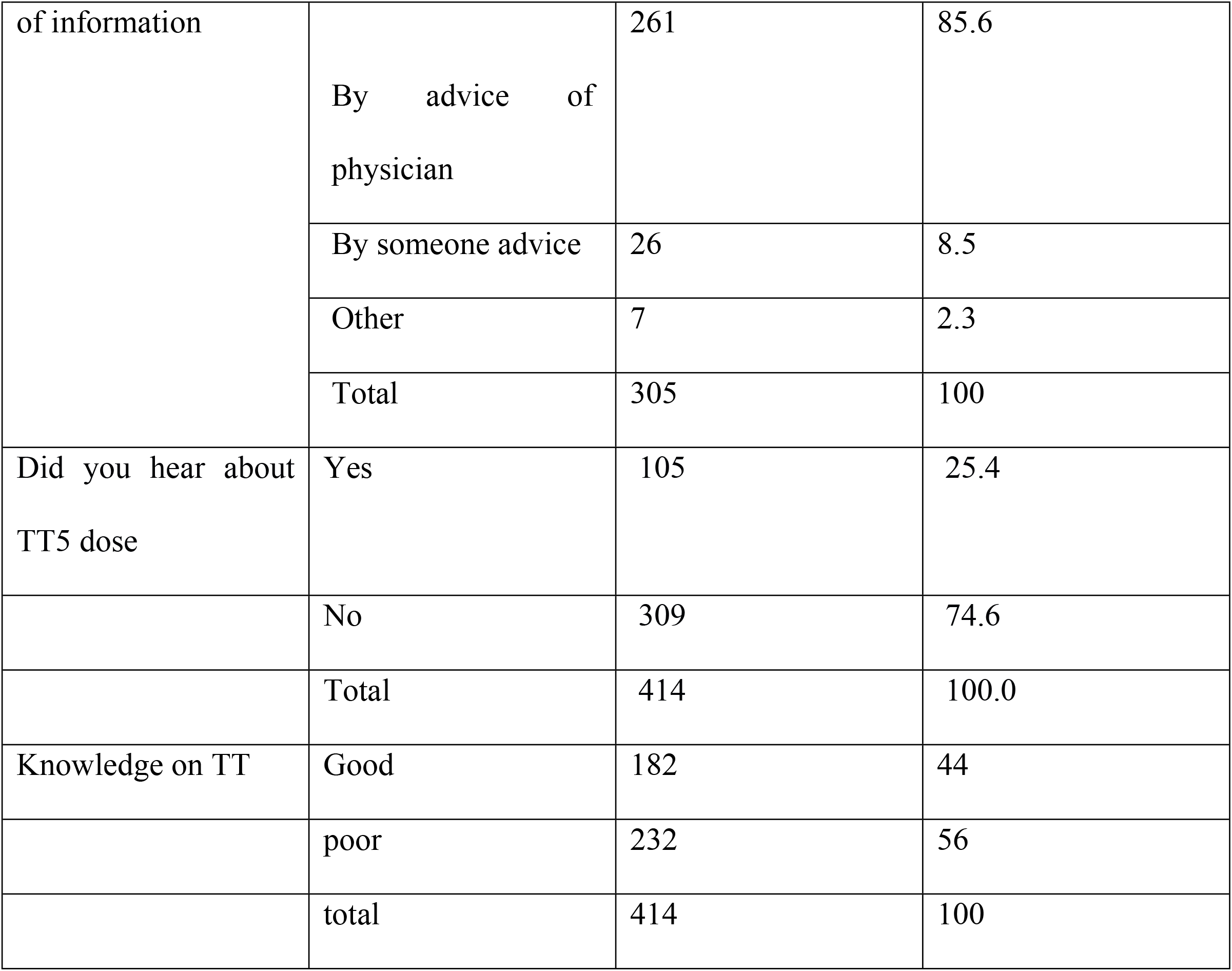
Knowledge and Uptake of TT immunization status, Haik town, South Wollo, Ethiopia, 2020.

Women reported that why they do not complete 5 consecutive dose according to EPI schedule is due to provider did not tell them next appointment and they did not know for extra dose 142(52%) and 87(31.9%) respectively (Figure 1).

**Figure 1.**
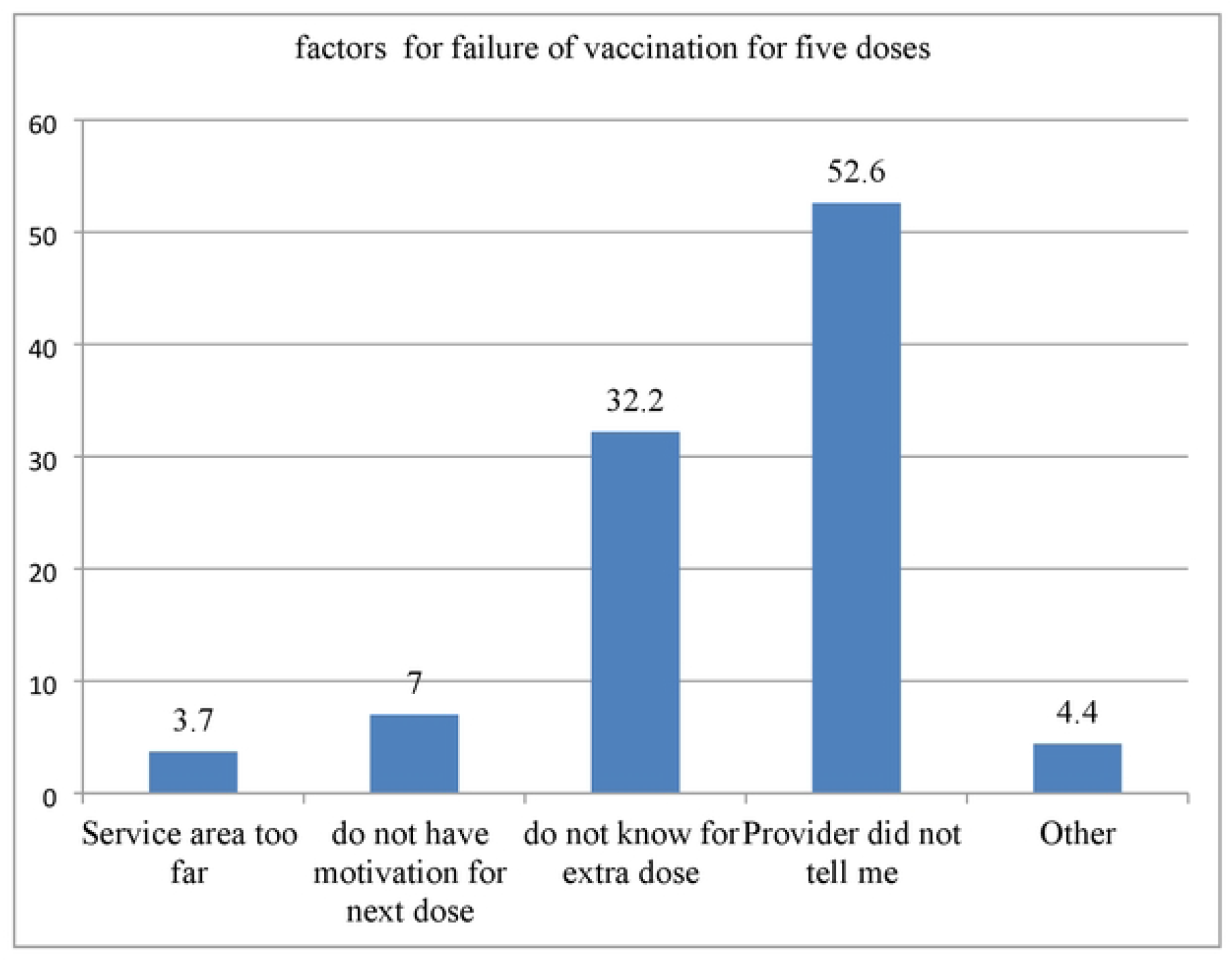
Reasons for failed complete TT vaccine, Haik town, South Wollo, Ethiopia, 2020

### Factors associated with TT vaccination

#### Factors associated with TT2 uptake

Variables with p-value less than 0.25 on bivariate analysis were entered to multivariable logistic analysis.Women who are educated secondary education and above, having history of ANC and having sort of information about TT5 were factors associated with TT vaccination.

Women who are educated secondary education and above (AOR=1.81, 95% CI: (1.10-2.97) were 1.81times more likely to vaccinate TT. Mothers who have ANC AOR =5.795%CI: (2.77-12.82) were six, times more likely to vaccinate TT. Women who have heard sort of information about TT5 AOR=5.7, CI **(2**.**53-12**.**86)**) 5.7 times more likely to take at least single dose of TTTable 4).

**Table 4.**
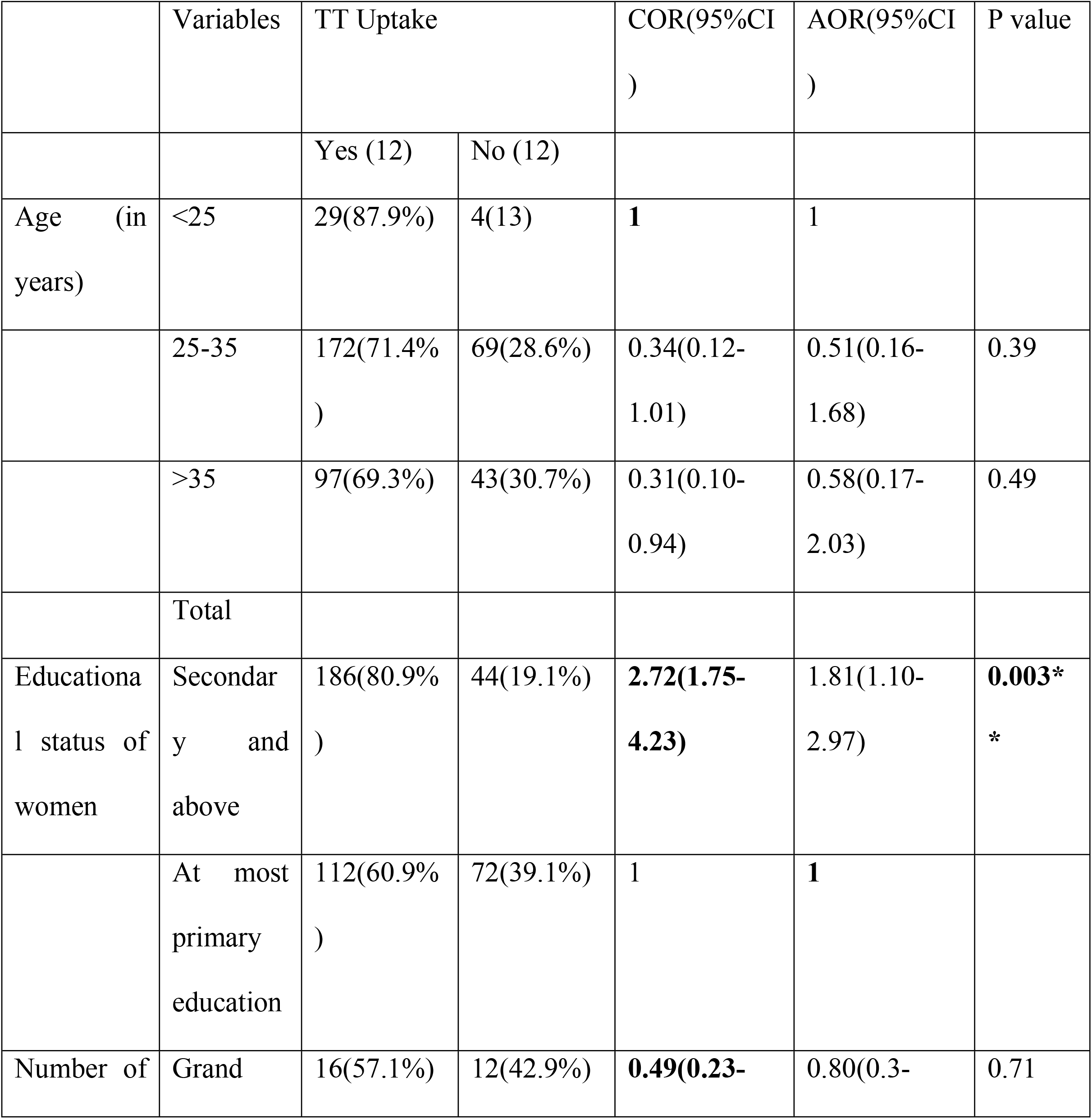
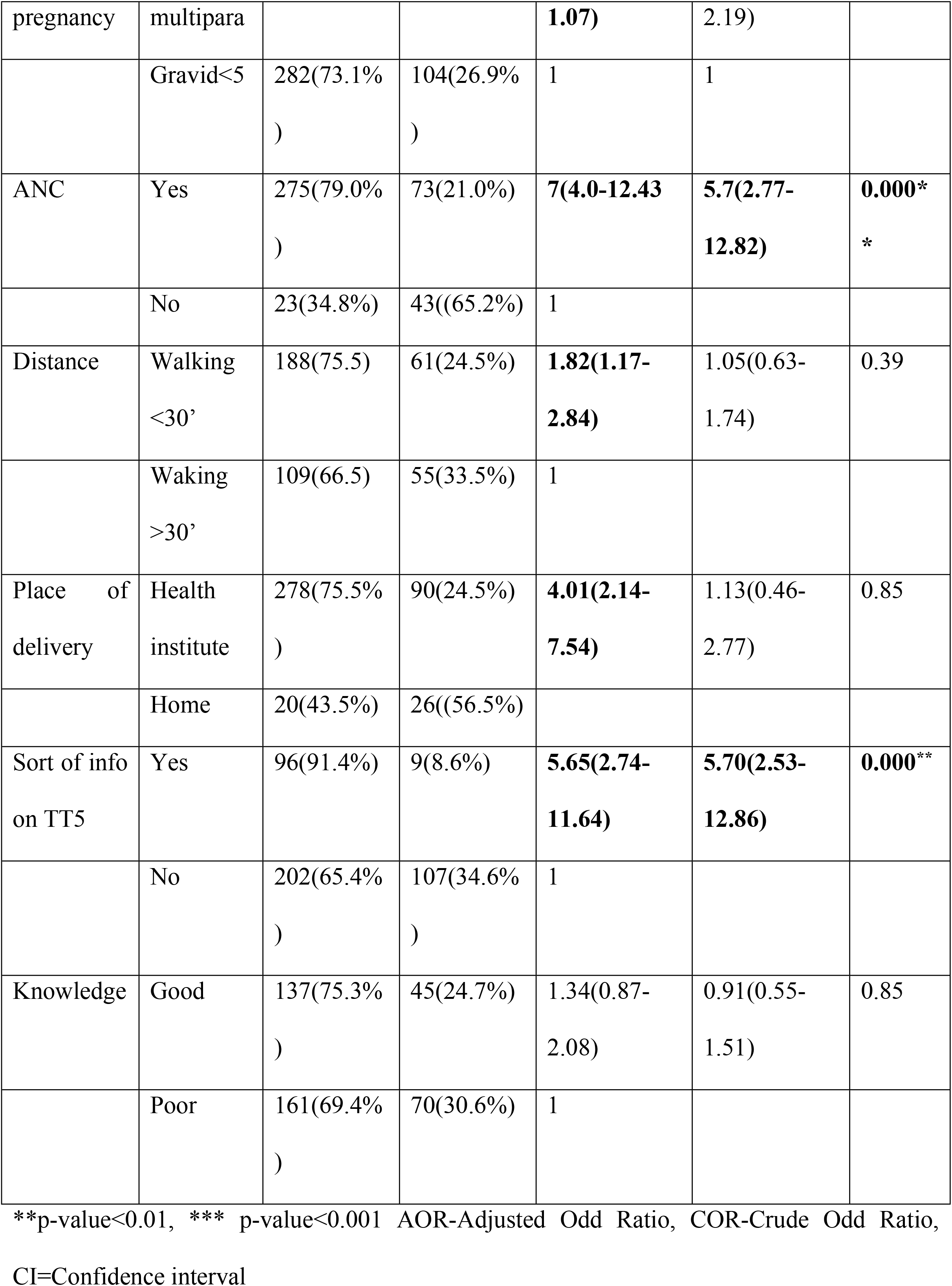
Multivariable analysis of knowledge and uptake of TT2 vaccine among Haik town, Southwollo, Ethiopia.

#### Factors associated with complete TT vaccine

Nearly half of 182(44%) of women in the reproductive age group had knowledge on complete TT5 immunization. Educational status, having sort of info on TT5 and knowledge about TT are significantly associated factors for full dose of TT according to EPI schedule. Women who are educated secondary education and above (AOR=**3**.**30**, 95% CI: ( **(1**.**11-9**.**77**) were **3**.**30** times more likely to vaccinate TT. Women who have any information about TT (AOR=**9**.**63**, 95% CI: **(3**.**87-23**.**96)**) were **9**.**63**times more likely to vaccinate TT. Mothers who have good knowledge on TT (AOR=**5**.**67**, 95% CI: **(3**.**04-15**.**75)**were **5**.**67**times more likely to vaccinate TT(Table 5)

**Table 5.**
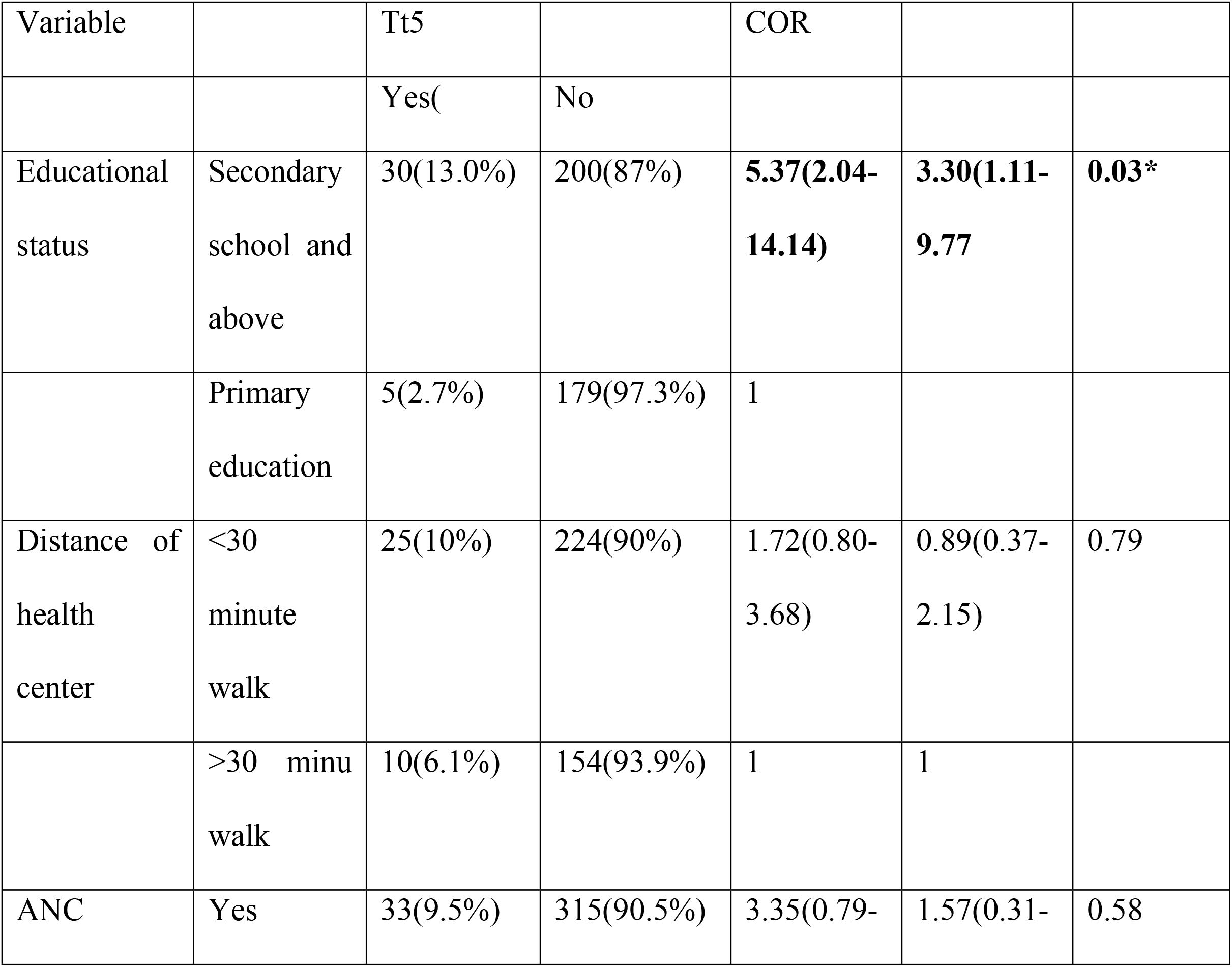
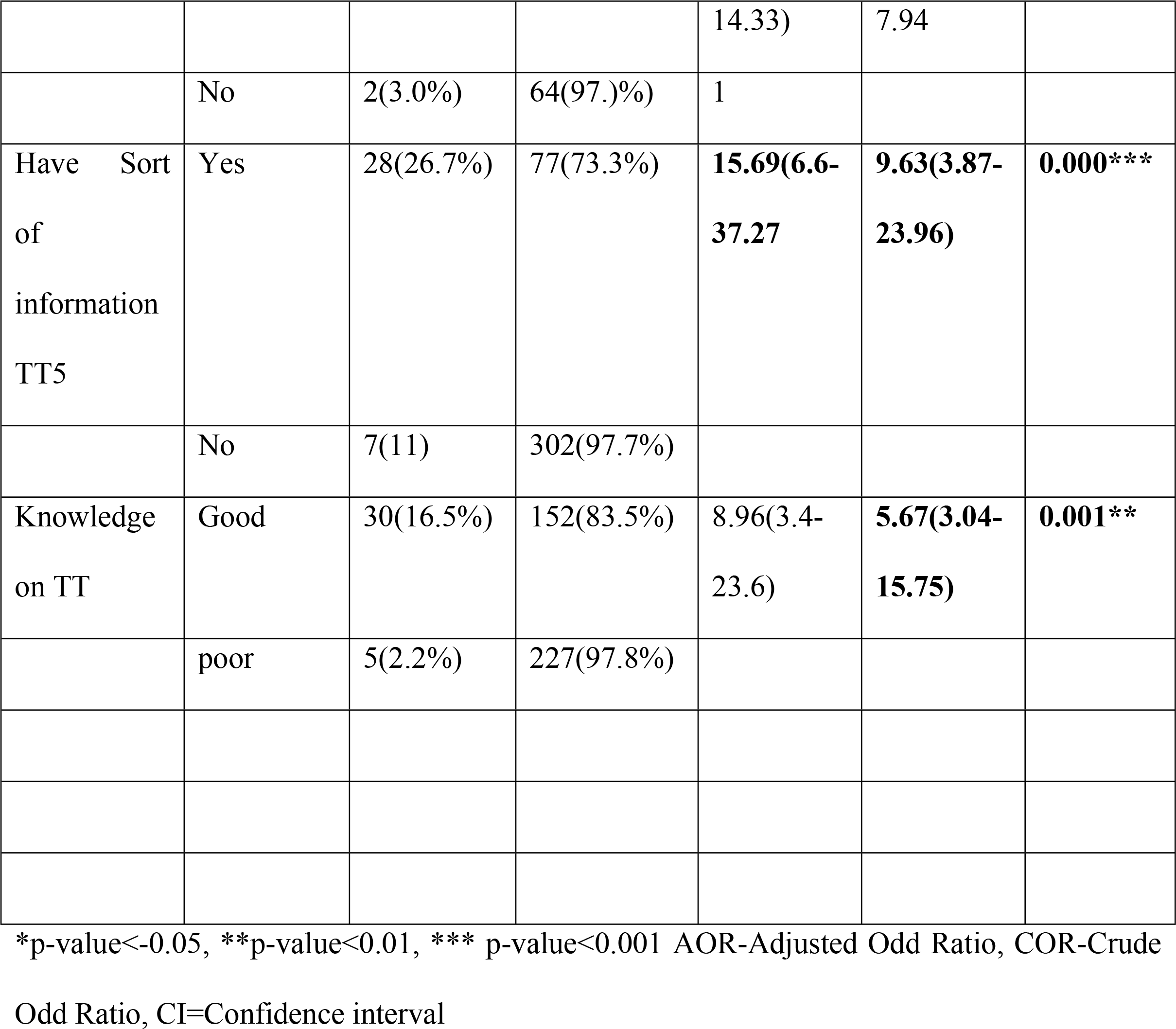
Multivariable analysis of knowledge and uptake of complete TT5 vaccine among Haik town, Southwollo, Ethiopia.

Thirty five (8.5%) of women of reproductive age group women completed TT5 according to EPI schedule with a 95% CI: 8.45-11.11%).

## Discussion

Magnitude of uptake of TT2 and above immunization at Hayk town was 71.2% at 95%CI 67.64%-76.32). But only 35(11.5%, valid dose, 8.5%, percent) had completed 5 doses of TT according to EPI schedule, the rest 270(88.5%) of women took TT when they are pregnant and not finish the full dose, they did not vaccinate after birth. One hundred nine (26.3%) had never take any dose of TT.

Magnitude of TT2 and above vaccination in this study is in line with the study conducted in Damboya, Kembata, (72.5%), at Dukem (65.1%) Debre Markos (12)

On the other hand the uptake of TT2 and above in this study is higher than the study conducted in, Errer district, Somali, Ethiopia which is 51.8%, Peshawar, Pakistan, 2010 which is 55.6%, in Dhaka, Bangladish,79%)(5, 11, 14). This may be due to most of pregnant women have at least two antenatal visit and hence they will take TT2 vaccine.

In this study complete dose of TT5 taken by women of reproductive age group is 35(11.5%), is lower than in Errer, Somali, 31(14.8%) of women took full dose TT and 33% in Dhaka Bangladesh, (5, 11). This may be due to poor knowledge of women about the disease as evidenced from the finding of this study, although a significant number of mothers knew about the vaccine.

In this study area mothers who have ANC AOR =5.795%CI: (2.77-12.82) were 5.8 times more likely to vaccinate TT. It is supported by the study in Hawzen, Tigrai described as women who attended one time antenatal care visits were 62% less likely to receive two doses of TT vaccine injection compared to mothers who attended four times and above antenatal care visits (15).

Women who attended secondary school and above were 1.81 times more likely to take TT vaccine (AOR=1.8: 95% CI:(1.10-2.97) than mothers who have educated less than high school education this study is in line with study in kembata, Dukem [AOR: 1.41, 95% CI: (1.84, 2.30)(12, 16)

In this study women who have some sort of information have 5.7 times more likely to vaccinate than those of who haven’t. This is inconsistent with study in Hawzen, Tigray women who got information from media were 4.5times more likely to take TT2 and above(15).

About 182(44%) of woman have good knowledge on TT vaccine. This is higher than the study done in Alexandria(8.7%)(17). This may be due to socio cultural difference.

## Conclusion

Fair proportions of women had uptake of TT2, but very low proportion of complete dose of TT5 according to expanded program on Immunization. Most of study participants have inadequate knowledge on TT vaccine Educational status, having ANC visit in the previous pregnancy and having sort of information on TT vaccination are significant association for the uptake of TT vaccine for women in the reproductive age group.

It is better to give emphasis prevention of diseases, adequate health education for full dose TT during ANC, and campaign vaccination of the community. Extra research should be done to explore the possible risk factors that hinder completion of the full dose vaccination.

## Data Availability

All relevant data are within the manuscript and its Supporting Information files.

## Supporting information

**S1 File. Questionnaire_Amharic_version**.

**S2 File. Questionnaire_English_version**.

**S1 Figure 1**

**S2 Figure 2**

**S1 Dataset**

## Acknowledgements

Our appreciation goes to Wollo university and all the women who took part in this study and for their patience.

## Author contributions

Conceptualization: Tiruset Gelaw,

Formal analysis: Tiruset Gelaw, Sindu Ayalew

Funding acquisition: Tiruset Gelaw,

Investigation: Tiruset Gelaw, Kassaw Beyene,

Methodology: Tiruset Gelaw, Sindu Ayalew, Kassa Beyene

Writing - original draft Tiruset Gelaw

Writing – review & editing: Tiruset Gelaw, Kassa Beyene, Sindu Ayalew

## Abbreviations

ANC: Antenatal care
MNT: maternal and neonatal sepsis
TT: tetanus toxoid
TTCV: tetanus toxoid containing vaccine
TTI: tetanus toxoid immunization

## Notes

### Competing Interest Statement

The authors have declared no competing interest.

### Funding Statement

The funders had no role in study design, data collection and analysis, decision to publish, or preparation of the manuscript.

### Author Declarations

Ethical clearance and permission was obtained from the Ethical review committee of Wollo University, college of medicine and health Science and the respective health institutions before the data collection process; cmhs/53/13/13

